# Modified Endoscopic Mucosal Resection Outperforms Endoscopic Submucosal Dissection for Rectal Neuroendocrine Tumors ≤10 mm: A Systematic Review and Meta Analysis

**DOI:** 10.64898/2026.02.10.26345872

**Authors:** Ke Pang, Lujing Ying, Hemiao Xu, Yabing Wang, Wangyang Chen, Daiyu Yang, Qiming Xiao, Shuai Li, Rutong Li, Hongwei Wang, Jingchao Gao, Peiyu Zhang, Jun Li, Kun He, Qiang Wang, Dong Wu

## Abstract

**Background:** Endoscopic resection is the standard treatment for rectal neuroendocrine tumors (r-NETs) ≤10 mm, yet the optimal technique remains controversial. Modified endoscopic mucosal resection (m-EMR) has emerged as a potential alternative compared to endoscopic submucosal dissection (ESD), but existing evidence is largely retrospective and the results of recent randomized controlled trials (RCTs) are inconclusive.

**Aims:** To compare the efficacy and safety of m-EMR versus ESD for r-NETs ≤10 mm.

**Methods:** We systematically searched CENTRAL, PubMed, Embase, and WanFang from January 1st, 1970 to December 23, 2025 for RCTs comparing m-EMR with ESD in r-NETs ≤10 mm. The GRADE framework assessed evidence certainty, while trial sequential analysis (TSA) controlled random errors and evaluated conclusion validity.

**Results:** Six RCTs involving 440 patients were analyzed. No significant difference between m-EMR and ESD was found in histologic complete resection (RR = 1.00, 95% CI 0.97–1.03; I^2^ = 0%), en bloc resection rates (P = 0.75) and procedure-related complications (P = 0.94). And m-EMR was associated with a significantly shorter procedure time (P<0.00001) and lower hospitalization cost (P<0.00001). The evidence was of moderate certainty; TSA confirmed its reliability, and both cumulative and sensitivity analyses supported the robustness.

**Conclusions:** Moderate-certainty evidence indicates m-EMR achieves oncologic outcomes comparable to ESD while offering clear advantages in procedural efficiency and cost for r-NETs ≤10 mm, supporting m-EMR possibly as a preferred endoscopic strategy in clinical practice.

## Introduction

Rectal neuroendocrine tumors (r-NETs) are rare subepithelial neoplasms that originate from neuroendocrine cells in the epithelial crypts, extending into the lamina propria or submucosa, while maintaining intact surface mucosa[1]. The increasing global incidence of r-NETs is largely attributed to the widespread use of screening colonoscopy, which has led to earlier and more frequent detection[2, 3]. The prognosis of r-NETs primarily depends on factors such as tumor size, endoscopic features (e.g., depression or ulceration), imaging findings (e.g., suspicious lymph nodes on endoscopic ultrasound [EUS] or magnetic resonance imaging [MRI]), and histopathological characteristics, including tumor grade and lymphangio-invasion[4, 5]. Tumors ≤ 10 mm are typically confined to the submucosal layer, presenting as small, sessile lesions with a relatively low risk of metastasis, resulting in a 100% long-term survival rate for patients without metastasis at diagnosis[6, 7]. This makes endoscopic resection the most appropriate curative option for such tumors.

The National Comprehensive Cancer Network (NCCN) guidelines recommend complete endoscopic resection as the first-line treatment for small (≤10 mm) incidental r-NETs with negative margins [8]. However, incomplete resection remains a significant clinical challenge, with reported incomplete resection rates ranging from 10% to 60%[9–12]. The optimal technique for endoscopic resection of r-NETs remains under debate. Given that r-NETs often invade the submucosa or deeper layers of the rectal wall, more advanced techniques such as endoscopic submucosal dissection (ESD) are recommended by guidelines compared to conventional endoscopic mucosal resection (c-EMR)[4, 13]. A meta-analysis demonstrated that ESD achieved a high complete resection rate of 89%, with adverse events occurring in 4% of cases and a local recurrence rate of <1%[14]. Furthermore, ESD outperforms c-EMR, showing a significantly higher complete resection rate (89% vs. 75%, p < 0.001). While ESD’s application has grown due to its superior R0 resection rate, its technical complexity, prolonged procedural time, and risk of adverse events are notable drawbacks[15].

Modified endoscopic mucosal resection (m-EMR) refers to a group of EMR-based techniques specifically designed to improve resection depth and margin status, particularly for submucosal tumors. The European Society of Gastrointestinal Endoscopy has also recommends m-EMR as the preferred treatment for r-NETs ≤10 mm[13], based on meta-analyses that revealed superior complete resection rates and procedural efficiency when compared with ESD[16, 17]. However, most studies included in these meta-analyses were retrospective, which may have introduced selection bias, as ESD might have been preferentially applied to more complex lesions, such as those with submucosal fibrosis or larger tumor size. M-EMR includes various techniques such as EMR with a ligation device (EMR-L)[18, 19], EMR using a transparent cap (EMR-C)[20], precut EMR (EMR-P)[21], anchored snare-tip EMR (ASEMR)[22], and simplified EMR (sEMR)[23], which omits submucosal injection. Recently, several randomized controlled trials (RCTs) have been published in this field, but their conclusions remain controversial[24–26].

Given the uncertainties and the lack of strong evidence in this field, we aimed to conduct a systematic review and meta-analysis of RCTs to synthesize the latest evidence comparing the efficacy and safety of m-EMR and ESD for rectal NETs ≤ 10 mm. In order to ensure the rigor of the results, the Grading of Recommendations Assessment, Development and Evaluation (GRADE) framework was used to assess the certainty of evidence[27], and trial sequential analysis (TSA) was used to control the risk of random errors and assess the conclusions[28].

## Methods

### Search Methodology

A literature search was conducted on Cochrane Central Register of Controlled Trials (CENTRAL), PubMed, Embase, and Wan Fang from January 1st, 1970 to December 23rd, 2025. No language or publication type restrictions were applied. Keywords used in this study mainly included “rectal neuroendocrine tumors”, “ESD” and “m-EMR” (detailed search strategies are listed in Supplementary File 1). All search results were saved to a citation management tool (EndNote), and the Bramer Method [29] was used to remove duplicates. In addition, the reference lists of all primary studies and review articles were manually searched. This meta-analysis was conducted following the Preferred Reporting Items for Systematic Reviews and Meta-Analyses (PRISMA) statement (Supplementary File 2) [30]. This study protocol was registered on PROSPERO (CRD420251275997).

### Study Eligibility

Eligible studies for inclusion in this meta-analysis were limited to RCTs comparing m-EMR and ESD in patients with histologically confirmed r-NETs ≤10 mm, in whom EUS and/or radiologic evaluation excluded muscularis propria invasion, lymph node involvement, and distant metastasis. The primary intervention was m-EMR, which includes techniques such as EMR-L, EMR-C, EMR-P, ASEMR, and sEMR, or other EMR modifications that meet the defined criteria. The comparator was conventional ESD. Studies conducted on animal models, editorials, abstracts, articles with incomplete data, and non-randomized studies were excluded. Two reviewers (LY and KP) independently screened the studies for eligibility and included potentially relevant articles. Full-text reviews were conducted based on the inclusion and exclusion criteria, and data were extracted by two reviewers (LY and HX). Any disagreements among reviewers were resolved through discussion or with the involvement of a third author (YW).

### Data extraction and quality assessment

Two authors (CW and DY) independently extracted data from eligible studies. The following information was collected: basic study details (year of publication and author names), study design characteristics (total duration of the study, number and location of study centers, allocation concealment, blinding procedures, withdrawals, and duration of follow-up), population characteristics (inclusion and exclusion criteria, number of participants, age, gender, presence of comorbidities, and lesion details), interventions (description of the m-EMR techniques used), and outcomes (identification of primary and secondary outcomes, time points at which outcomes were reported, complications related to resection, and therapeutic costs). Any discrepancies between the reviewers were resolved by discussion or by consulting a third author (KP).

### Study quality assessment

Two review authors (KH and KP) independently assessed risk of bias of included studies according to the following seven domains: random sequence generation, allocation concealment, blinding of participants and personnel, blinding of outcome assessment, incomplete outcome data, selective outcome reporting and other possible bias. We graded the risk of bias for each domain as high, low, or unclear and provided information from the study report together with a justification for our judgement in the ‘Risk of bias’ tables.

### Outcomes evaluated

The primary outcome of this study is histological complete resection (R0) at the initial endoscopy. This outcome is defined as complete en bloc resection of the targeted r-NET, with both horizontal and vertical free margins, confirmed by histopathological examination.

Important secondary outcomes include en bloc resection, procedure time, hospitalization cost and procedure-related complications, which are assessed for level of certainty of evidence. Procedure-related complications were evaluated as a composite outcome, including procedural bleeding, delayed bleeding, and postoperative complications. Procedural bleeding referred to bleeding events occurring during the endoscopic procedure that required additional hemostatic intervention. Delayed bleeding was defined as bleeding occurring within 30 days after the index procedure, resulting in a hemoglobin decrease of >2 g/dL or necessitating additional endoscopic, radiological, or surgical intervention. Postoperative complications included adverse events occurring after resection, such as bleeding, perforation, or infection. Other secondary outcomes include intraoperative and postoperative bleeding, negative vertical and lateral margin and length of hospital stay.

Sensitivity analysis of primary outcome was performed using the leave-one-out method to assess the robustness of pooled results by sequentially omitting each individual study and recalculating the combined effect estimates.

## Statistical Analysis

### Measures of treatment effect and heterogeneity

Meta-analysis was conducted by using RevMan version 5.4.1 (Cochrane Training, London, United Kingdom). A random-effects model was employed to obtain pooled data, and the DerSimonian and Laird method was used to estimate the between-study variance. For dichotomous data, effect sizes were calculated as risk ratios (RRs) with 95% CIs. For continuous data, we extracted the mean value and standard deviation (SD). To calculate effect sizes, we utilized mean differences (MDs) with 95% confidence intervals (CIs). In instances where data were reported as medians, minimum and maximum values, and/or first and third quartiles, we applied a data transformation method developed by Wan et al. to convert the data into mean values and SDs to ensure consistency in pooling the results[31]. Additionally, since a random-effects model was used for pooling, prediction intervals were also computed using the approach proposed by Higgins et al[32].

The heterogeneity of the studies was assessed using Cochran’s Q test based on inverse variance weights and by calculating the I^2^ statistic. An I^2^ statistic > 50% or Cochran’s Q test p < 0.1 was considered moderate to substantial heterogeneity. In cases where the I^2^ statistic exceeded 80% (indicating substantial heterogeneity), we provided a visual representation of the individual study effects rather than conducted a meta-analysis[33].

### Assessment of risk of bias and level of certainty of evidence

For each study, two review authors (KP and LY) independently assessed the risk of bias using criteria outlined in the Cochrane Collaboration Risk of Bias tool (RoB 2)[34]. The tool evaluates five domains: randomization bias, deviations from intended interventions, missing outcome data bias, outcome measurement bias, and selective reporting bias. Each study was rated as “low risk,” “some concerns,” or “high risk”. To evaluate the certainty of the evidence for the primary outcome and important secondary outcomes, we utilized the Grading of Recommendations Assessment, Development, and Evaluation (GRADE) framework. GRADE Pro version 3.6 software from McMaster University, Hamilton, Ontario, Canada (http://gradepro.org/), was employed. No publication bias assessment was performed as there were fewer than 10 studies included and the power of the tests for funnel plot was low[27].

The certainty of evidence for the primary and important secondary outcomes was assessed using the Grading of Recommendations Assessment, Development, and Evaluation (GRADE) framework[35]. Evidence certainty was rated as high, moderate, low, or very low. Any disagreements in evidence grading were resolved through discussion among the review team.

### Assessment of evidence robustness and sufficiency

Sensitivity analyses were conducted to assess the influence of individual studies on the pooled estimates. Each study was sequentially excluded from the meta-analysis, and the pooled effect size was recalculated to evaluate the stability of the overall conclusions.

In addition, a cumulative meta-analysis was performed for the primary outcome based on the chronological order of publication. Trials were added one at a time according to their publication year, and pooled effect estimates were recalculated after each addition. This method was used to explore the temporal stability of the pooled effect, assuming that improvements in endoscopic techniques and operator experience over time might influence the outcomes.

Trial sequential analysis (TSA) was used to control the risk of random errors caused by low sample sizes and repeated significance testing of included studies and adjust the thresholds for statistical significance in the meta-analysis[28]. TSA was performed via TSA software version 0.9 beta (Copenhagen Trial Unit, Centre for Clinical Intervention Research, Copenhagen, Denmark)[36]. For discontinuous data, we set effect measure ‘RR’ and model as ‘Random-effects (DL)’ in TSA software. The required sample size for primary outcome was calculated based on anticipated relative risk reduction of included RCTs with low-risk bias, with a power (1-β) of 80% and type I errorαof 5%. Other data were calculated based on average incidence in all included RCTs. Heterogeneity correction was autogenerated by the software based on model variance. The statistical significance of the meta-analysis was evaluated according to the position of the cumulative Z curve with conventional boundary, trial sequential monitoring boundary (TSMB), and futility boundary in the figure. A stable and firm conclusion was reached if the cumulative Z curve crossed the trial sequential monitoring boundary or entered the futility area below the futility boundary[37]. All statistical analyses were performed using Stata software (version 12.0; StataCorp, College Station, TX, USA) or R software.

## Results

### Screening and characterization of studies

After conducting an initial search of four databases (CENTRAL, PubMed, Embase, and Wan Fang), a total of 54 records were identified. Out of these, 16 duplicate studies were removed. Subsequently, 32 articles were excluded during the screening process as they did not meet the predetermined selection criteria. Finally, six randomized controlled trials (RCTs) were included in this meta-analysis. The PRISMA flow diagram is shown in **Figure 1**.

**Figure 1.**
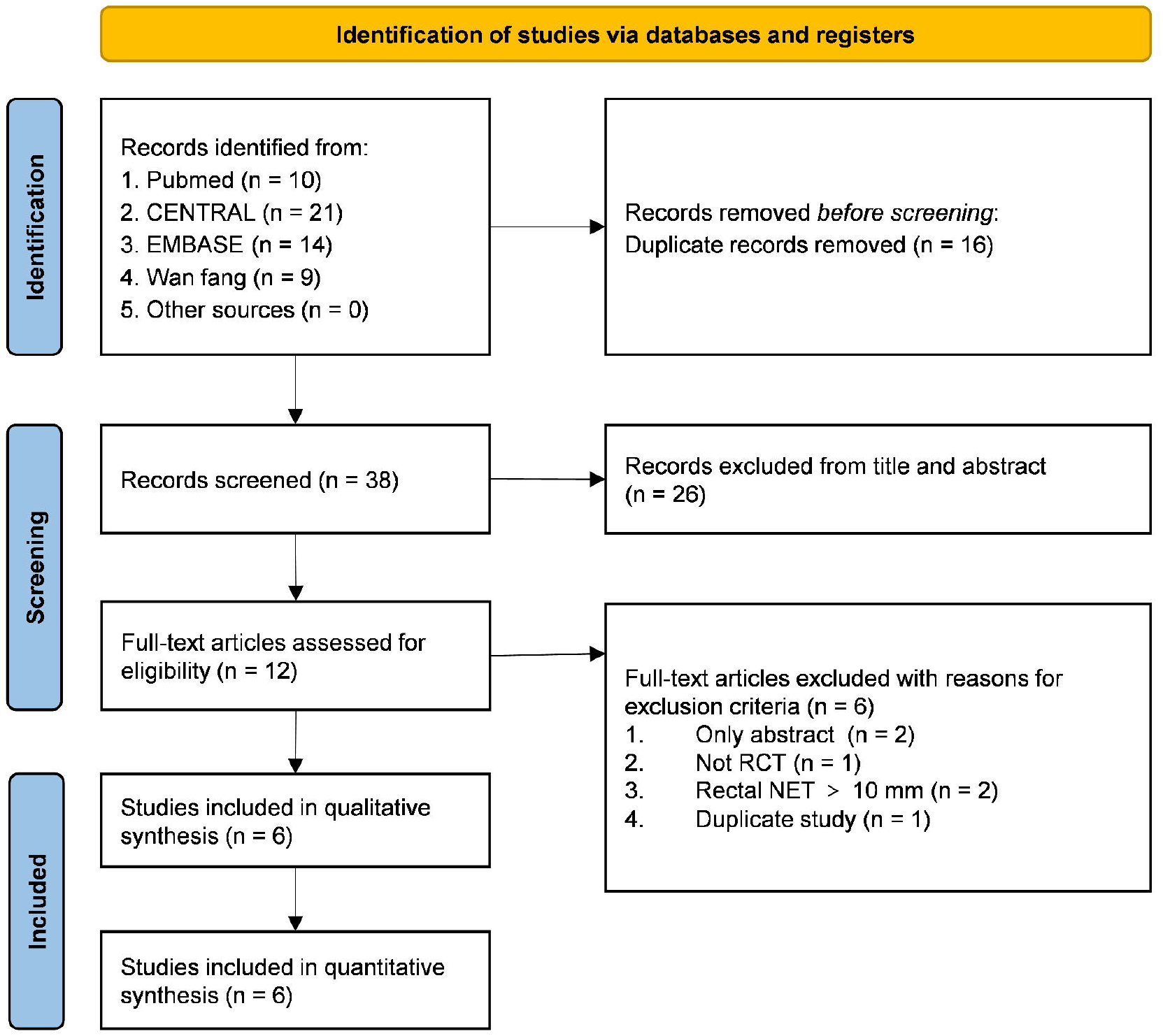
PRISMA flowchart represents the flow of information through the different phases of the systematic review and meta-analysis. PRISMA: Preferred Reporting Items for Systematic Reviews and Meta-Analyses.

Table 1. shows the details of the included studies. A total of 440 participants were enrolled in this meta-analysis, with 214 participants treated with m-EMR and the remaining 226 participants with ESD. Sensitivity analysis showed exceptional robustness of the findings. Sequential omission of individual trials yielded consistent pooled RRs ranging from 0.96 to 1.05, with all P-values exceeding 0.70 (range: 0.7388–0.9483). Importantly, heterogeneity remained at zero (I^2^ = 0%, Tau^2^ = 0, Tau = 0) regardless of which study was excluded, confirming that no single trial disproportionately influenced the overall conclusion. The narrow confidence intervals (95% CI: 0.97–1.04) persisted across all iterations, indicating high precision and stability of the pooled estimate.

**Table 1.**
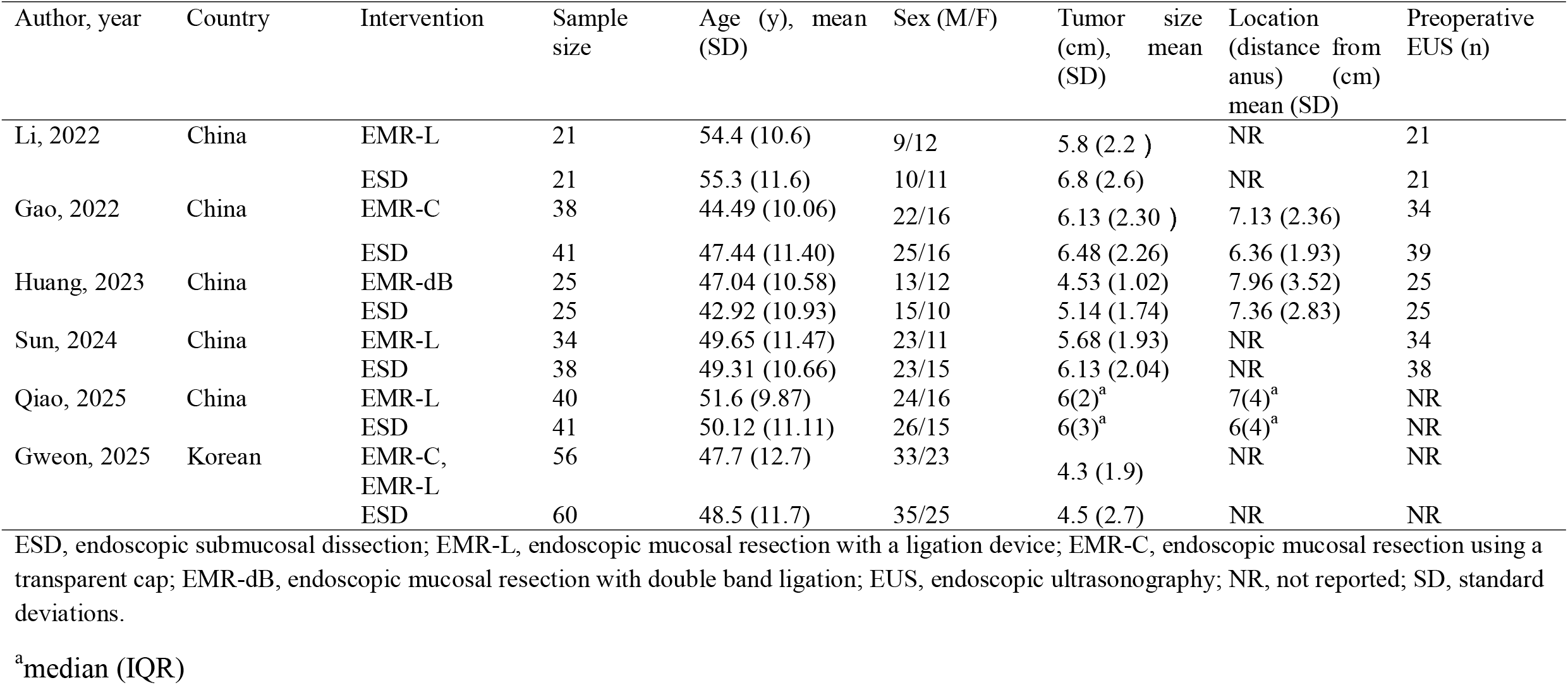
Study characteristics.

### Risk of bias in the included studies and certainty of evidence for outcomes

The risk of bias in the included RCTs was assessed using the Cochrane RoB 2.0 tool. Four studies (Li et al. 2022, Gao et al. 2022, Qiao et al. 2025, and Gweon et al. 2025) showed low overall risk, with proper randomization, no deviations, complete outcome data, unbiased measurements, and no selective reporting. The other two (Huang et al. 2023, Sun et al. 2024) had some concerns due to unclear randomization but were otherwise low risk. No studies had a high risk (**Figure 2**).

**Figure 2.**
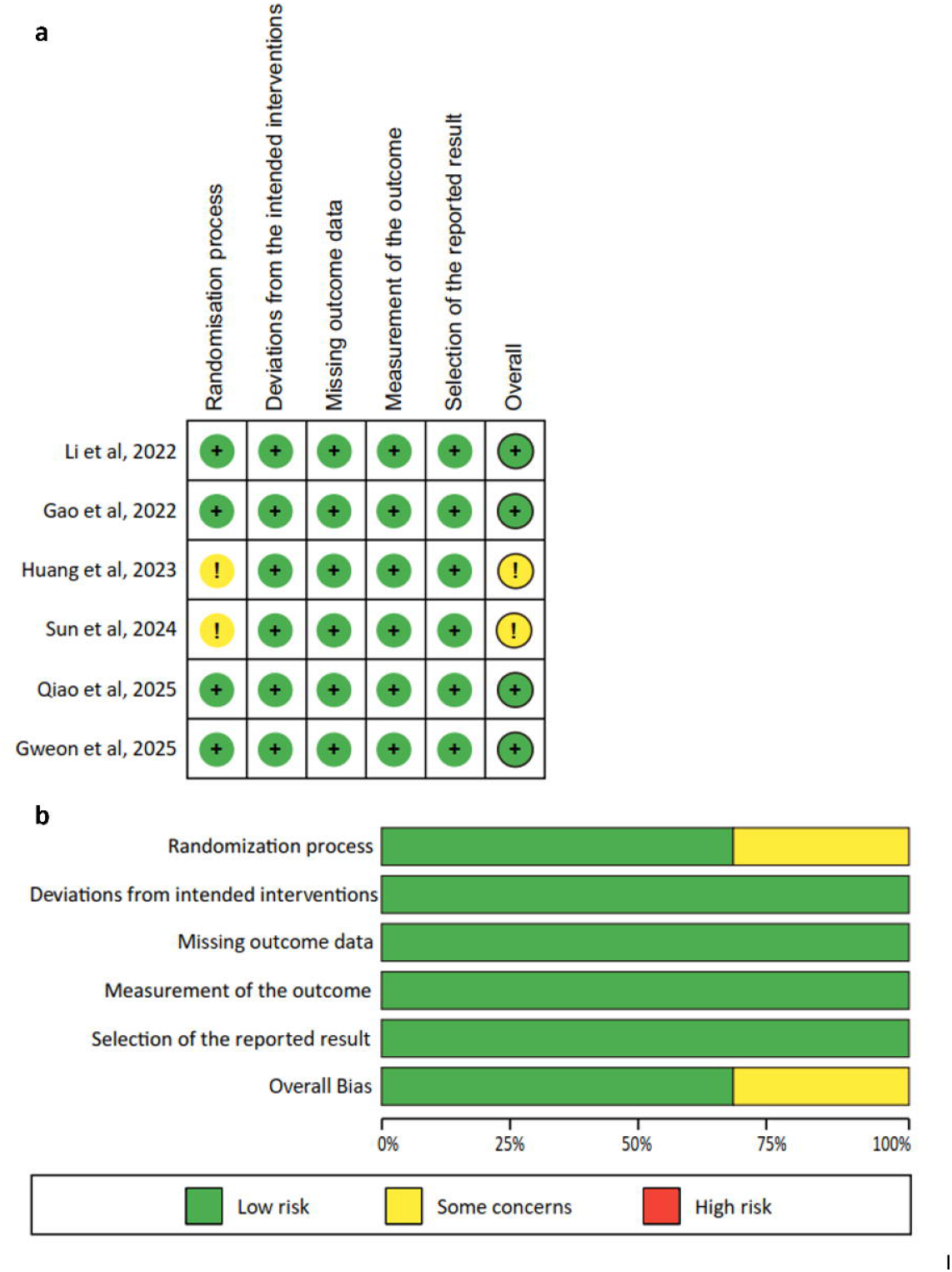
Methodological quality of included studies according to the revised Cochrane tool to assess risk of bias (RoB 2). (A) Risk of bias summary. (B) Risk of bias graph.

We assessed the quality of evidence for the main outcomes mentioned above using the GRADE methodology which was shown in Table 2.

**Table 2.** Summary of main findings.

| Outcomes | Anticipated absolute effects* (95%CI) |  | Relative effect (95%CI) | No. of patients (studies) | Certainty of the evidence (GRADE) |
| --- | --- | --- | --- | --- | --- |
|  | Risk with ESD | Risk with m-EMR |  |  |  |
| Histologic complete resection | 925 per 1,000 | 925 per 1,000 (897 to 953) | RR 1.00 (0.97 to 1.03) | 440 (6 RCTs) | ⊕⊕⊕⊖ Moderate<sup>a</sup> |
| En Bloc Resection | 991 per 1,000 | 991 per 1,000 (961 to 1,000) | RR 1.00 (0.97 to 1.02) | 440 (6 RCTs) | ⊕⊕⊕⊖ Moderate<sup>a</sup> |
| Procedure-related complications | 21 per 1,000 | 17 per 1,000 (3 to 122) | RR 0.92 (0.12 to 5.74) | 368 (5 RCTs) | ⊕⊕⊕⊖ Moderate<sup>a</sup> |
| Procedure time | The mean procedure time was 23.64 minutes | MD 14.93 minutes lower (17.14 lower to 12.71 lower) | - | 276 (3 RCTs) | ⊕⊕⊕⊖ Moderate<sup>a</sup> |
| Hospitalization cost | The mean hospitalization cost was 3144.68 dollars | MD 745.63 dollars lower (917.14 lower to 574.12 lower) | - | 151 (2 RCTs) | ⊕⊕⊕⊖ Moderate<sup>a</sup> |

GRADE Working Group grades of evidence. High Certainty: further research is very unlikely to change our confidence in the estimate of effect; Moderate Certainty: further research is likely to have an important impact on our confidence in the estimate of effect and may change the estimate; Low Certainty: further research is very likely to have an important impact on our confidence in the estimate of effect and is likely to change the estimate; Very low quality: we are very uncertain about the estimate; CI confidence interval; RR Relative risk; \*The basis for the assumed risk is the average control group proportion across all comparison; m-EMR: modified endoscopic mucosal resection; ESD: endoscopic submucosal dissection; <sup>a</sup>: downgraded one level for imprecision.

### Results of the meta-analysis Primary outcome

#### Histologic complete resection

All six RCTs assessed the histologic complete resection. Meta-analysis revealed that there was no significant difference between m-EMR and ESD (RR: 1.00, 95% CI: 0.97–1.03; P =0.95; I^2^ = 0%; n = 440; RCTs = 6; moderate quality of evidence) (**Figure 3A**). Leave-one-out sensitivity analysis demonstrated stable pooled estimates for histologic complete resection (RR range: 0.96–1.05), with consistently non-significant P-values and minimal heterogeneity (I^2^ = 0%), indicating that no individual trial influenced the overall conclusion (**Figure 3B**). Cumulative meta-analysis further showed that the pooled effect estimate remained consistency as trials were added in chronological order, with no change in effect direction, statistical significance, or heterogeneity, indicating a stable and robust absence of difference between m-EMR and ESD (Figure 3C). TSA revealed that the required information size was 700. The cumulative Z curve entered the futility area below the futility boundary, suggesting that the negative results were reliable (**Figure 3D**).

**Figure 3.**
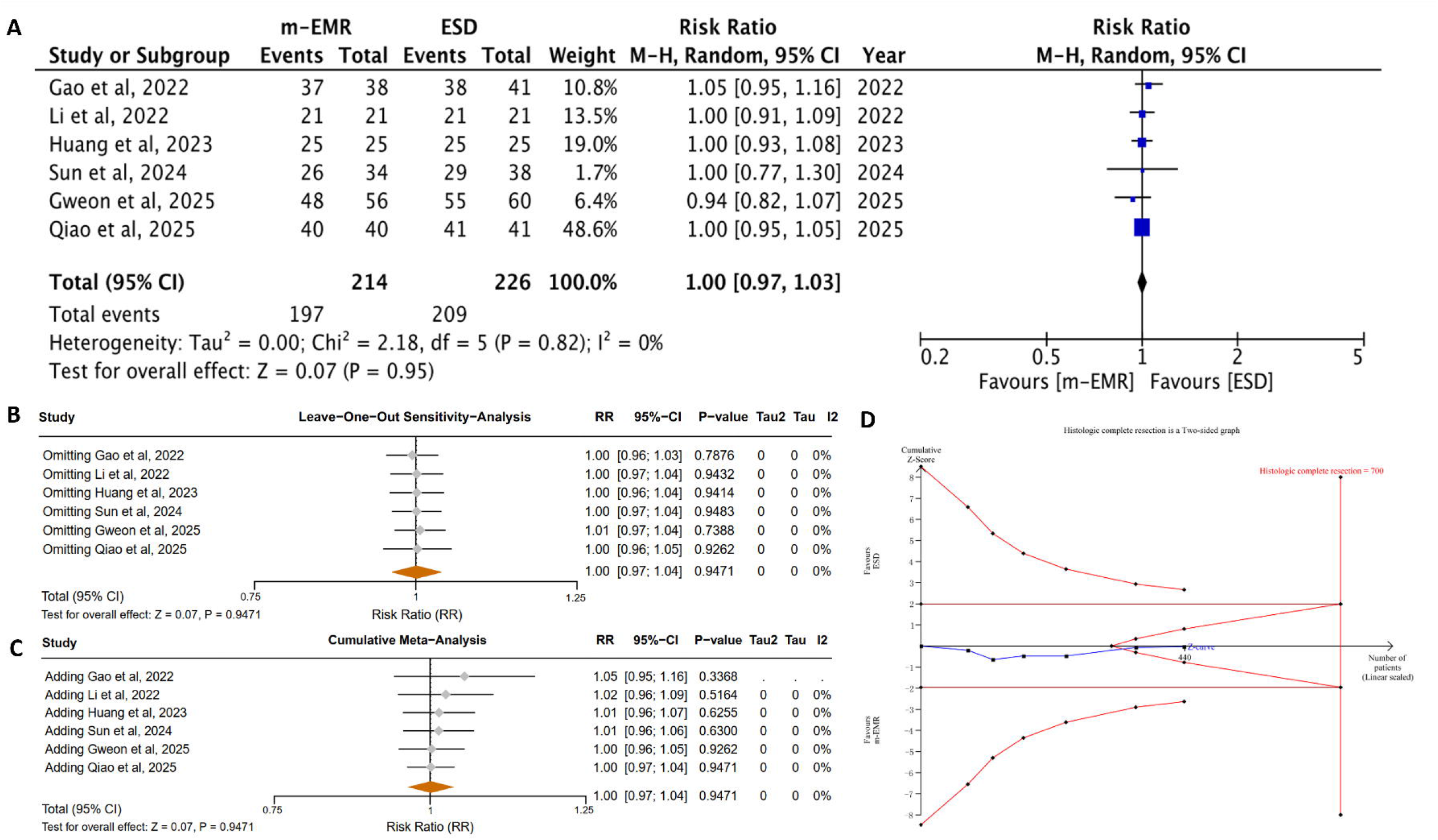
Forest plots and TSA illustrating the primary outcome. (A) Forest plots for histologic complete resection; (B) TSA for histologic complete resection; (C) Sensitivity analysis for histologic complete resection; (D) Cumulative analysis for histologic complete resection.

### Important secondary outcomes

#### en bloc resection

There was no significant difference in en bloc resection between patients treated with m-EMR and ESD (RR: 1.00, 95% CI: 0.97-1.02; P = 0.75; I^2^ = 0%; n = 440; RCTs = 6; moderate certainty of evidence) (**Figure 4A**). TSA revealed that the required information size was 465. The cumulative Z curve entered the futility area below the futility boundary, suggesting that the negative results were reliable (**Figure 4B**).

**Figure 4.**
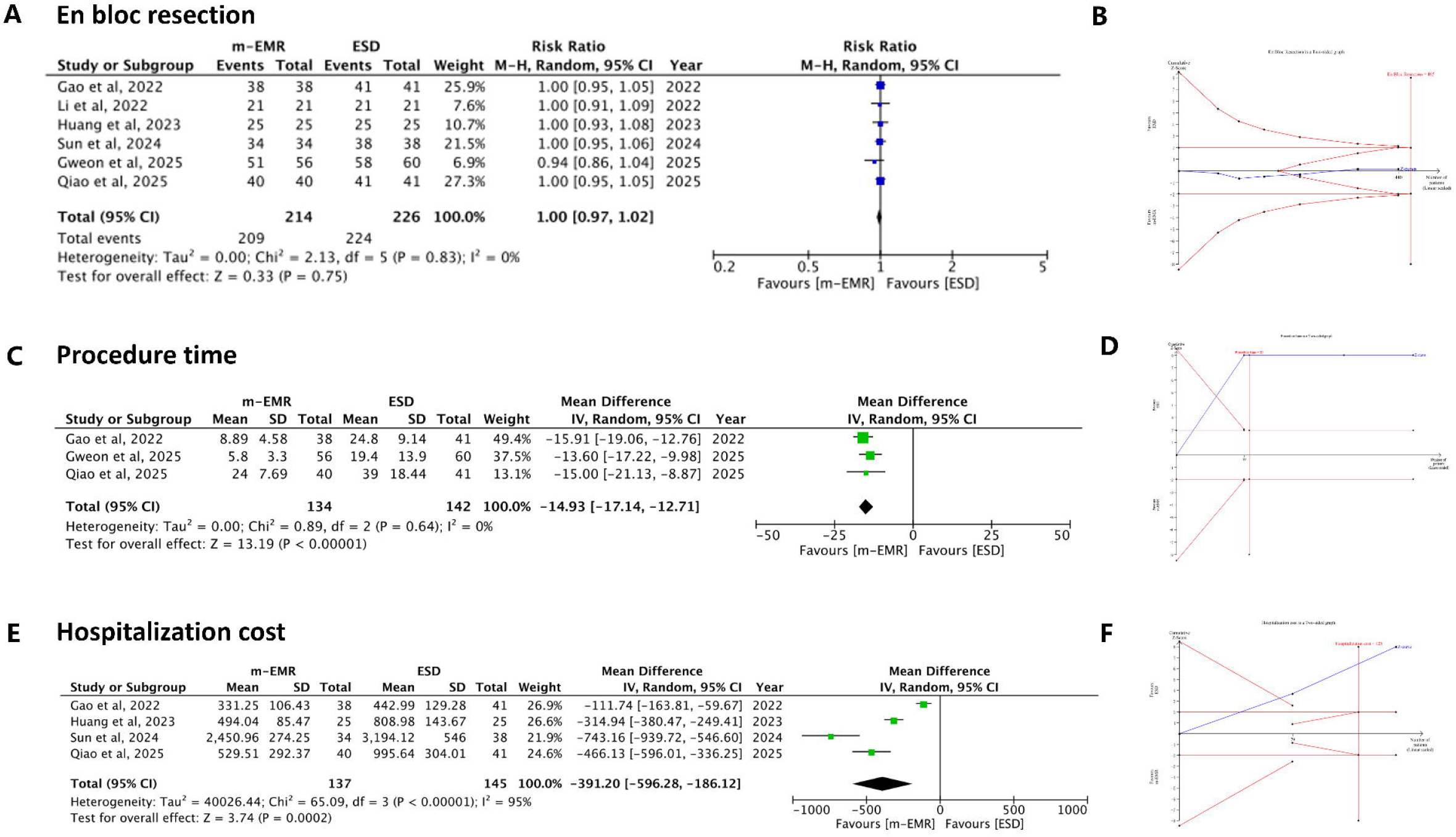
Forest plots and TSA for secondary outcomes(A, B)Forest plots and TSA for en bloc resection; (C,D) Forest plots and TSA for procedure time; (E,F) Forest plots and TSA for hospitalization cost. m-EMR, modified-endoscopic mucosal resection; ESD, endoscopic submucosal dissection; CI, confidence interval, TSA, Trial sequential analysis.

#### Procedure time

Meta-analysis of three RCTs (Gao et al. 2022; Gweon et al. 2025; Qiao et al. 2025) revealed that m-EMR led to a significantly shorter procedure time than ESD (MD = -14.93, 95% CI: -17.14 – -12.17; P <0.00001; I^2^ = 0%; n = 276; RCTs = 3; moderate certainty of evidence) (**Figure 4C**). The other three RCTs were excluded due to a lack of a clear definition of the procedure time. TSA revealed that the required information size was 85. The cumulative Z curve for procedure time crossed the TSMB and required sample size, suggesting the result was reliable (**Figure 4D**).

#### Hospitalization cost

Meta-analysis of two RCTs with clear definition (Gao et al. 2022; Sun et al. 2024) revealed that the hospitalization cost was significantly lower in the m-EMR group than in the ESD group (MD: -745.63, 95% CI: -917.14– -574.12; P<0.00001; I^2^ = 0%; n = 151; RCTs = 2; moderate quality of evidence) (**Figure 4E**). It was noted that m-EMR was also associated with significantly lower costs than ESD in the other two excluded RCTs (Huang et al. 2023; Qiao et al. 2025). TSA revealed that the required information size was 125. The cumulative Z curve for hospitalization cost crossed the TSMB and the required information size, suggesting the result was reliable (**Figure 4F**).

#### Procedure-related complication

Meta-analysis revealed no significant difference in procedure-related complications between patients treated with m-EMR and ESD (RR = 0.92, 95% CI: 0.09-8.86; P = 0.94; I^2^ = 48%; n = 368; RCTs = 5; moderate certainty of evidence) (**Figure 5A**).

**Figure 5.**
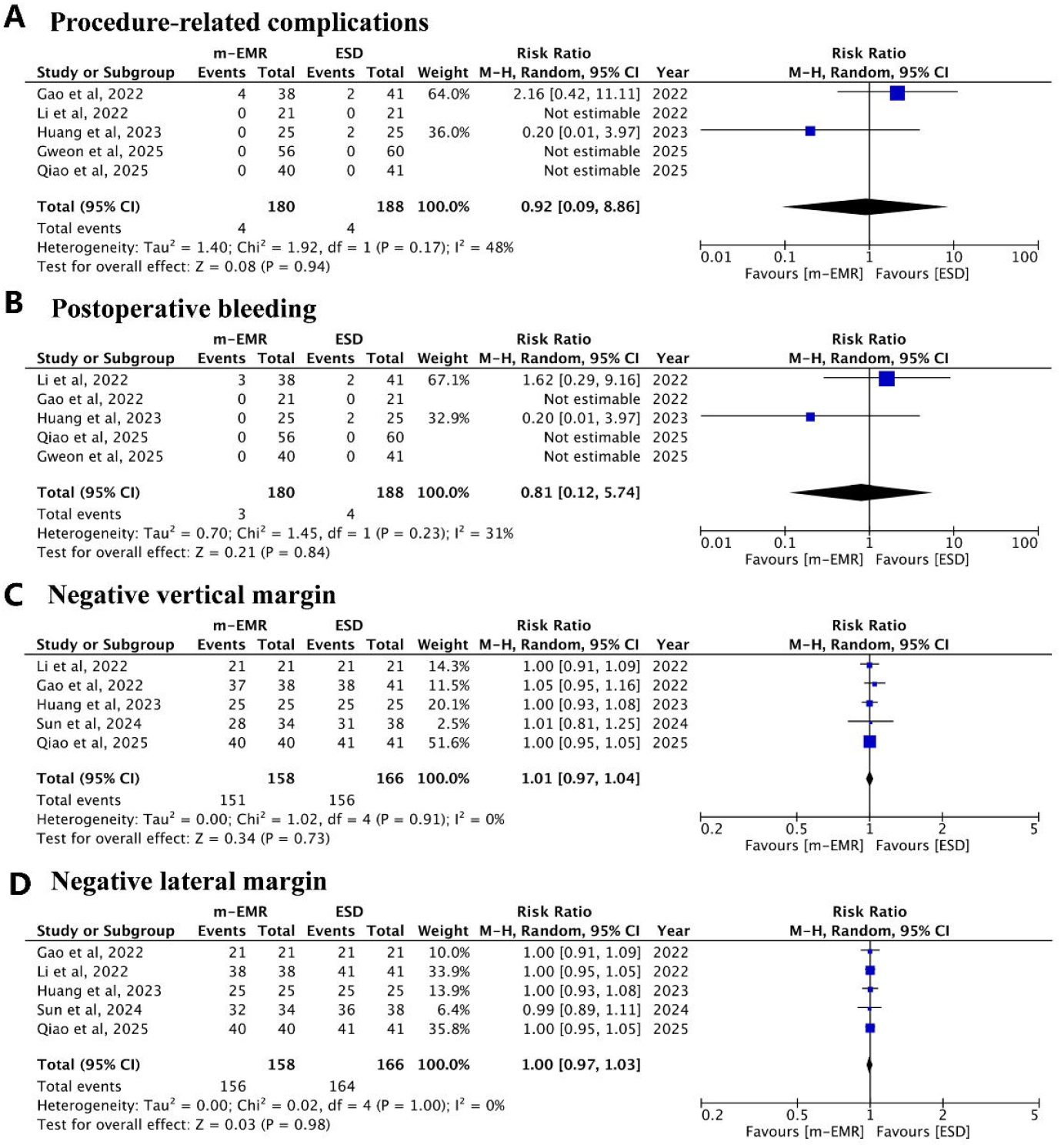
Forest plots of other outcomes. (A) Forest plots for procedure-related complications; (B) Forest plots for postoperative bleeding; (C) Forest plots for negative vertical margin; (D) Forest plots for negative lateral margin.

### Other secondary outcomes

#### Intraoperative and postoperative bleeding

Five RCTs (n=368) clearly recorded the intraoperative and postoperative bleeding with a specific definition. Only Gweon et al. 2025 reported one case of uncontrollable intraoperative bleeding in the ESD group that could not be managed via endoscopic interventions. Meta-analysis revealed there was no significant difference in postoperative bleeding between m-EMR and ESD (RR: 0.81, 95% CI: 0.12-5.74; P = 0.84;I^2^ = 31%; n = 368; RCTs = 5) (**Figure 5B**).

#### Negative vertical and lateral margin

Meta-analysis revealed that there was no significant difference between m-EMR and ESD in negative vertical margin (RR: 1.01, 95% CI: 0.97-1.04; P = 0.73; I^2^ = 0%; n = 324; RCTs = 5), and negative lateral margin (RR: 1.00, 95% CI: 0.97-1.03; P = 0.98; I^2^ = 0%; n = 324; RCTs = 5) (**Figure 5C**).

#### Hospital stay

Two RCTs reported the length of hospital stay. Meta-analysis was not performed due to the high heterogeneity (I^2^ = 84%). We presented the individual results of these two studies in Table 3. Gao et al. 2022 found that there was no significant difference in length of hospital stay between m-EMR and ESD (6.24 vs 5.73 days, P=0.427), while Sun et al. 2024 reported a significantly shorter length of hospitalization in m-EMR compared with ESD (4 vs 5 days, P = 0.001) (**Figure 5D**).

## Discussion

Endoscopic resection is currently recommended as the standard treatment for r-NETs measuring ≤10 mm[4, 8, 13]; however, the optimal endoscopic technique remains a matter of debate, largely owing to the rapid evolution of endoscopic technologies and the paucity of high-quality comparative evidence. Although ESD has demonstrated superior oncologic outcomes compared with c-EMR, particularly given the high rate of positive vertical margins associated with c-EMR (reported to be up to 50%)[4], its technical complexity, longer procedural time, and higher procedural demands have limited its widespread adoption in routine clinical practice[38]. In response, several modified EMR (m-EMR) techniques have been developed to enhance lesion capture and resection depth, thereby improving the likelihood of achieving histologic complete resection[21, 24–26, 39–44].

In this meta-analysis of RCTs, m-EMR demonstrated oncologic efficacy comparable to that of ESD, with no significant differences observed in histologic complete resection, en bloc resection, or margin-negative resection rates. Importantly, modified EMR was associated with significantly shorter procedure times and substantially lower hospitalization costs, supporting its overall procedural efficiency advantage over ESD in the treatment of rectal NETs ≤10 mm. The certainty of evidence for these key outcomes was graded as moderate, and TSA confirmed that the accrued randomized evidence was sufficient to support these conclusions.

It should be emphasized that m-EMR represents a broad category of techniques rather than a single standardized procedure. In this study five of the six RCTs exclusively used EMR-L[24–26, 39] or EMR-C[26, 41], with Huang et al. further exploring a double-band ligation technique[40]. Despite procedural variations, these techniques share a common axial-traction mechanism designed to maximize resection depth and vertical margin clearance. By aspirating the lesion and surrounding submucosal tissue into a ligation device or transparent cap before snare resection, m-EMR facilitates deeper vertical excision without the need for meticulous submucosal dissection. Although ESD allows precise control of the dissection plane, endoscopists often adopt a relatively shallow submucosal layer to minimize the risk of muscularis propria injury and perforation, which may limit achievable resection depth. Consequently, for small r-NETs ≤10 mm that are not constrained by cap or ligation device diameter, m-EMR can achieve histologic outcomes comparable to ESD while offering superior procedural efficiency.

Beyond oncologic equivalence, procedural efficiency represents a critical consideration in real-world practice. In this analysis, m-EMR was associated with significantly shorter procedure times than ESD, reflecting a simplified workflow that reduces operator fatigue and endoscopy suite utilization. In addition, material costs were consistently lower with m-EMR due to reduced reliance on specialized accessories such as electrosurgical knives and submucosal injection devices. Collectively, these advantages support m-EMR as a highly efficient and cost-effective strategy for r-NETs ≤10 mm, particularly in high-volume or resource-limited settings.

Compared with prior meta-analyses[17, 45, 46], the present study reached a more conservative and methodologically robust conclusion by restricting inclusion to RCTs and incorporating TSA, sensitivity analysis and cumulative analysis. Earlier syntheses predominantly pooled retrospective or non-randomized studies, which are inherently susceptible to selection bias, where ESD may have been preferentially applied to more complex lesions, such as those with submucosal fibrosis or larger tumor size, while modified EMR was selected for technically favorable cases. In addition, TSA demonstrated that the available RCT evidence was adequate to support conclusions of comparable oncologic efficacy between the two techniques, while leave-one-out sensitivity analyses and cumulative meta-analysis confirmed the stability of non-inferiority [47].

Several limitations should be acknowledged. First, although only RCTs were included, the overall sample size remained modest. Nevertheless, the reliability of the primary outcomes was reinforced by TSA, which confirmed that the accumulated evidence was sufficient to support the main conclusions. Second, although m-EMR represents a broad category, the six included RCTs predominantly evaluated EMR-L and EMR-C. However, it is noteworthy that other m-EMR variants, such as EMR-P[21], ASEMR[22], sEMR[23], have demonstrated comparable efficacy and safety to ESD in non-randomized studies, providing supportive evidence that reinforces the overall conclusions of this meta-analysis. Third, subgroup analyses according to lesion location, endoscopist experience, or institutional volume were not performed due to limited reporting and the small number of eligible trials. Fourth, most included studies were conducted in East Asian populations, which may introduce a degree of regional bias and limit the generalizability of the findings. Therefore, although the results appear robust and consistent, further large-scale, multicenter randomized studies in diverse populations are warranted. Finally, long-term outcomes, including local recurrence and disease-specific survival, were insufficiently reported, precluding assessment beyond histologic complete resection.

## Conclusion

In summary, based on evidence from RCTs, and supported by sensitivity, cumulative analysis, and trial sequential analysis confirming the robustness and reliability of the findings, moderate-certainty evidence suggests that m-EMR achieves oncologic outcomes comparable to ESD while offering clear advantages in procedural efficiency and cost, supporting its preferential use for r-NETs ≤10 mm in routine clinical practice. Nevertheless, given that most available evidence originates from East Asian populations, large-scale, multicenter randomized studies are still warranted to confirm the generalizability of these findings in broader real-world settings.

## Data Availability

Not applicable.

## Acknowledgements

The authors would like to thank Tao Wang, for his invaluable advice in the preparation of this manuscript.

## Reference

1. Dabkowski K, Szczepkowski M, Kos-Kudła B, Starzynska T. Endoscopic management of rectal neuroendocrine tumours. How to avoid a mistake and what to do when one is made? Endokrynol Pol. 2020;71:343–9. 10.5603/EP.a2020.0045.

2. Scherübl H. Rectal carcinoids are on the rise: early detection by screening endoscopy. Endoscopy. 2009;41:162–5. 10.1055/s-0028-1119456.

3. Dasari A, Shen C, Halperin D, Zhao B, Zhou S, Xu Y, et al. Trends in the Incidence, Prevalence, and Survival Outcomes in Patients With Neuroendocrine Tumors in the United States. JAMA Oncol. 2017;3:1335–42. 10.1001/jamaoncol.2017.0589.

4. Panzuto F, Parodi MC, Esposito G, Massironi S, Fantin A, Cannizzaro R, et al. Endoscopic management of gastric, duodenal and rectal NETs: Position paper from the Italian Association for Neuroendocrine Tumors (Itanet), Italian Society of Gastroenterology (SIGE), Italian Society of Digestive Endoscopy (SIED). Dig Liver Dis. 2024;56:589–600. 10.1016/j.dld.2023.12.015.

5. Moon CM, Huh KC, Jung S-A, Park DI, Kim WH, Jung HM, et al. Long-Term Clinical Outcomes of Rectal Neuroendocrine Tumors According to the Pathologic Status After Initial Endoscopic Resection: A KASID Multicenter Study. Am J Gastroenterol. 2016;111:1276–85. 10.1038/ajg.2016.267.

6. Fahy BN, Tang LH, Klimstra D, Wong WD, Guillem JG, Paty PB, et al. Carcinoid of the rectum risk stratification (CaRRs): a strategy for preoperative outcome assessment. Ann Surg Oncol. 2007;14:1735–43. 10.1245/s10434-006-9311-6.

7. Ngamruengphong S, Kamal A, Akshintala V, Hajiyeva G, Hanada Y, Chen Y-I, et al. Prevalence of metastasis and survival of 788 patients with T1 rectal carcinoid tumors. Gastrointest Endosc. 2019;89:602–6. 10.1016/j.gie.2018.11.010.

8. Shah MH, Goldner WS, Benson AB, Bergsland E, Blaszkowsky LS, Brock P, et al. Neuroendocrine and Adrenal Tumors, Version 2.2021, NCCN Clinical Practice Guidelines in Oncology. J Natl Compr Cancer Netw JNCCN. 2021;19:839–68. 10.6004/jnccn.2021.0032.

9. Zhuang X, Zhang S, Chen G, Luo Z, Hu H, Huang W, et al. Risk factors and clinical outcomes of incomplete endoscopic resection of small rectal neuroendocrine tumors in southern China: a 9-year data analysis. Gastroenterol Rep. 2023;11:goac084. 10.1093/gastro/goac084.

10. Fine C, Roquin G, Terrebonne E, Lecomte T, Coriat R, Do Cao C, et al. Endoscopic management of 345 small rectal neuroendocrine tumours: A national study from the French group of endocrine tumours (GTE). United Eur Gastroenterol J. 2019;7:1102–12. 10.1177/2050640619861883.

11. Matsuno K, Miyamoto H, Kitada H, Yoshimatsu S, Tamura F, Sakurai K, et al. Comparison of endoscopic submucosal resection with ligation and endoscopic submucosal dissection for small rectal neuroendocrine tumors: A multicenter retrospective study. DEN Open. 2023;3:e163. 10.1002/deo2.163.

12. Cha B, Shin J, Ko WJ, Kwon KS, Kim H. Prognosis of incompletely resected small rectal neuroendocrine tumor using endoscope without additional treatment. BMC Gastroenterol. 2022;22:293. 10.1186/s12876-022-02365-z.

13. Rinke A, Ambrosini V, Dromain C, Garcia-Carbonero R, Haji A, Koumarianou A, et al. European Neuroendocrine Tumor Society (ENETS) 2023 guidance paper for colorectal neuroendocrine tumours. J Neuroendocrinol. 2023;35:e13309. 10.1111/jne.13309.

14. de Mestier L, Brixi H, Gincul R, Ponchon T, Cadiot G. Updating the management of patients with rectal neuroendocrine tumors. Endoscopy. 2013;45:1039–46. 10.1055/s-0033-1344794.

15. Cao Y, Liao C, Tan A, Gao Y, Mo Z, Gao F. Meta-analysis of endoscopic submucosal dissection versus endoscopic mucosal resection for tumors of the gastrointestinal tract. Endoscopy. 2009;41:751–7. 10.1055/s-0029-1215053.

16. Pan J, Zhang X, Shi Y, Pei Q. Endoscopic mucosal resection with suction vs. endoscopic submucosal dissection for small rectal neuroendocrine tumors: a meta-analysis. Scand J Gastroenterol. 2018;53:1139–45. 10.1080/00365521.2018.1498120.

17. Zhou C, Zhang F, We Y. Efficacy of endoscopic mucosal resection versus endoscopic submucosal dissection for rectal neuroendocrine tumors ≤10mm: a systematic review and meta-analysis. Ann Saudi Med. 2023;43:179–95. 10.5144/0256-4947.2023.179.

18. Ono A, Fujii T, Saito Y, Matsuda T, Lee DT y, Gotoda T, et al. Endoscopic submucosal resection of rectal carcinoid tumors with a ligation device. Gastrointest Endosc. 2003;57:583–7. 10.1067/mge.2003.142.

19. Kim HH, Park SJ, Lee SH, Park HU, Song CS, Park MI, et al. Efficacy of endoscopic submucosal resection with a ligation device for removing small rectal carcinoid tumor compared with endoscopic mucosal resection: analysis of 100 cases. Dig Endosc Off J Jpn Gastroenterol Endosc Soc. 2012;24:159–63. 10.1111/j.1443-1661.2011.01190.x.

20. Park SB, Kim HW, Kang DH, Choi CW, Kim SJ, Nam HS. Advantage of endoscopic mucosal resection with a cap for rectal neuroendocrine tumors. World J Gastroenterol. 2015;21:9387–93. 10.3748/wjg.v21.i31.9387.

21. So H, Yoo SH, Han S, Kim G-U, Seo M, Hwang SW, et al. Efficacy of Precut Endoscopic Mucosal Resection for Treatment of Rectal Neuroendocrine Tumors. Clin Endosc. 2017;50:585–91. 10.5946/ce.2017.039.

22. Kim J, Kim J, Oh EH, Ham NS, Hwang SW, Park SH, et al. Anchoring the snare tip is a feasible endoscopic mucosal resection method for small rectal neuroendocrine tumors. Sci Rep. 2021;11:12918. 10.1038/s41598-021-92462-y.

23. Zou L, Zou L, Yang Y, Zhou W, Wu X, Guo T, et al. Safety and Efficacy of Simplified EMR Versus ESD for Rectal Neuroendocrine Tumors ≤ 10 Mm: A Retrospective Cohort Study. J Clin Med. 2025;14:6125. 10.3390/jcm14176125.

24. Qiao Wen, Guo Yudong, Tang Xiufen, Tang Lixin, Xi Tengfei, Qi Zijuan. Comparison of two endoscopic resection methods for rectal neuroendocrine neoplasm≤10 mm: A prospective randomized controlled study. Chin J Gastrointest Endosc Electron Eidition. 2025;12:177–83. 10.3877/cma.j.issn.2095-7157.2025.03.003.

25. Sun T. EMR and ESD in the treatment of rectal neuroendocrine tumors: a randomized controlled trial. [R735.37]. Shandong First Medical University; 2024.

26. Gweon T-G, Kim SJ, Myung DS, Jung Y, Lee J, Yang D-H. Optimal endoscopic treatment for rectal neuroendocrine tumors, confirmed after forceps biopsy: A Korean Association for the Study of Intestinal Diseases multicenter, randomized non-inferiority trial. Gastrointest Endosc. 2025;:S0016-5107(25)02211-4. 10.1016/j.gie.2025.11.035.

27. Higgins JPT, Thomas J, Chandler J, Cumpston M, Li T, Page MJ, et al. Cochrane handbook for systematic reviews of interventions version 6.4 (updated August 2023). Cochrane, 2023. Available Train Cochrane Orghandbook. 2023.

28. Brok J, Thorlund K, Wetterslev J, Gluud C. Apparently conclusive meta-analyses may be inconclusive--Trial sequential analysis adjustment of random error risk due to repetitive testing of accumulating data in apparently conclusive neonatal meta-analyses. Int J Epidemiol. 2009;38:287–98. 10.1093/ije/dyn188.

29. Bramer WM, Giustini D, de Jonge GB, Holland L, Bekhuis T. De-duplication of database search results for systematic reviews in EndNote. J Med Libr Assoc JMLA. 2016;104:240–3. 10.3163/1536-5050.104.3.014.

30. Page MJ, McKenzie JE, Bossuyt PM, Boutron I, Hoffmann TC, Mulrow CD, et al. The PRISMA 2020 statement: an updated guideline for reporting systematic reviews. BMJ. 2021;372:n71. 10.1136/bmj.n71.

31. Wan X, Wang W, Liu J, Tong T. Estimating the sample mean and standard deviation from the sample size, median, range and/or interquartile range. BMC Med Res Methodol. 2014;14:135. 10.1186/1471-2288-14-135.

32. Higgins JPT, Thompson SG, Spiegelhalter DJ. A re-evaluation of random-effects meta-analysis. J R Stat Soc Ser A Stat Soc. 2009;172:137–59. 10.1111/j.1467-985X.2008.00552.x.

33. Higgins JPT, Altman DG, Gøtzsche PC, Jüni P, Moher D, Oxman AD, et al. The Cochrane Collaboration’s tool for assessing risk of bias in randomised trials. BMJ. 2011;343:d5928. 10.1136/bmj.d5928.

34. Sterne JAC, Savović J, Page MJ, Elbers RG, Blencowe NS, Boutron I, et al. RoB 2: a revised tool for assessing risk of bias in randomised trials. BMJ. 2019;366:4898. 10.1136/bmj.l4898.

35. Balshem H, Helfand M, Schünemann HJ, Oxman AD, Kunz R, Brozek J, et al. GRADE guidelines: 3. Rating the quality of evidence. J Clin Epidemiol. 2011;64:401–6. 10.1016/j.jclinepi.2010.07.015.

36. Thorlund K, Engstrøm J, Wetterslev J, Brok J, Imberger G, Gluud C. User manual for trial sequential analysis (TSA) Copenhagen trial unit, centre for clinical intervention research. Cph Den. 2011;1:1–115.

37. Wetterslev J, Jakobsen JC, Gluud C. Trial Sequential Analysis in systematic reviews with meta-analysis. BMC Med Res Methodol. 2017;17:39. 10.1186/s12874-017-0315-7.

38. Chen Q, Yuan Y, Linghu E. Advances in the diagnosis and treatment of gastrointestinal tumors under the concept of super minimally invasive surgery. J Transl Intern Med. 2025;13:183–6. 10.1515/jtim-2025-0023.

39. Li Weiguang, Sun Yunwei, Sun Jing, Zhang Benya, Wang Huafeng, Qian Aihua. Management of rectal neurosecretory tumors less than 1 cm in diameter: comparison of two endoscopic procedures. Theory Pract Intern Med. 2022;17:289–94. 10.16138/j.1673-6087.2022.04.004.

40. Huang J-L, Gan R-Y, Chen Z-H, Gao R-Y, Li D-F, Wang L-S, et al. Endoscopic mucosal resection with double band ligation versus endoscopic submucosal dissection for small rectal neuroendocrine tumors. World J Gastrointest Surg. 2023;15:440–9. 10.4240/wjgs.v15.i3.440.

41. Gao X, Huang S, Wang Y, Peng Q, Li W, Zou Y, et al. Modified Cap-Assisted Endoscopic Mucosal Resection Versus Endoscopic Submucosal Dissection for the Treatment of Rectal Neuroendocrine Tumors ≤10 mm: A Randomized Noninferiority Trial. Am J Gastroenterol. 2022;117:1982–9. 10.14309/ajg.0000000000001914.

42. Taghiakbari M, Kim DHD, Djinbachian R, von Renteln D. Endoscopic resection of large non-pedunculated colorectal polyps: current standards of treatment. eGastroenterology. 2024;2:e100025. 10.1136/egastro-2023-100025.

43. Kim J, Kim J, Oh EH, Ham NS, Hwang SW, Park SH, et al. Anchoring the snare tip is a feasible endoscopic mucosal resection method for small rectal neuroendocrine tumors. Sci Rep. 2021;11:12918. 10.1038/s41598-021-92462-y.

44. Li D, Xie J, Hong D, Liu G, Wang R, Jiang C, et al. Efficacy and safety of ligation-assisted endoscopic submucosal resection combined with endoscopic ultrasonography for treatment of rectal neuroendocrine tumors. Scand J Gastroenterol. 2022;57:734–9. 10.1080/00365521.2022.2033828.

45. Kitagawa Y, Suzuki T, Miyakawa A, Okimoto K, Matsumura T, Shiratori T, et al. Comparison of endoscopic submucosal dissection and modified endoscopic mucosal resection for rectal neuroendocrine tumors. Sci Rep. 2025;15:5424. 10.1038/s41598-024-82082-7.

46. Chen J, Ye J, Zheng X, Chen J. Endoscopic treatments for rectal neuroendocrine tumors: a systematic review and network meta-analysis. J Gastrointest Surg Off J Soc Surg Aliment Tract. 2024;28:301–8. 10.1016/j.gassur.2023.12.016.

47. Zhang J, He C. Evidence-based rehabilitation medicine: definition, foundation, practice and development. Med Rev 2021. 2024;4:42–54. 10.1515/mr-2023-0027.

